# The impact of three progressively introduced interventions on second wave daily COVID-19 case numbers in Melbourne, Australia

**DOI:** 10.1101/2021.05.29.21258055

**Authors:** Allan Saul, Nick Scott, Tim Spelman, Brendan S. Crabb, Margaret Hellard

## Abstract

**Background:** The city of Melbourne, Australia experienced two waves of the COVID-19 epidemic peaking, the first in March and a more substantial wave in July 2020. During the second wave, a series of control measure were progressively introduced that initially slowed the growth of the epidemic then resulted in decreasing cases until there was no detectable local transmission.

**Methods:** To determine the relative efficacy of the progressively introduced intervention measures, we modelled the second wave as a series of exponential growth and decay curves. We used a linear regression of the log of daily cases vs time, using a four-segment linear spline model corresponding to implementation of the three successive major public health measures. The primary model used all reported cases between 14 June and 15 September then compared the projection of the model with observed cases predict future case trajectory up until the 31 October to assess the use of exponential models in projecting the future course and planning future interventions. The main outcome measures were the exponential daily growth constants, analysis of residuals and estimates of the 95% confidence intervals for the expected case distributions, comparison of predicted daily cases.

**Results:** The exponential growth/decay constants in the primary analysis were: 0.122 (s.e. 0.004), 0.035 (s.e. 0.005), -0.037 (s.e. 0.011), and -0.069 (s.e. 0.003) for the initial growth rate, Stage 3, stage 3 + compulsory masks and Stage 4, respectively. Extrapolation of the regression model from the 14 September to the 31 October matched the decline in observed cases over this period.

**Conclusions:** The four-segment exponential model provided an excellent fit of the observed reported case data and predicted the day-to-day range of expected cases. The extrapolated regression accurately predicted the decline leading to epidemic control in Melbourne.

## Background

Australia experienced a rise in coronavirus disease (COVID-19) cases in early 2020, peaking on 28 March 2020 and then declining in April after federal and state governments introduced strict community controls, travel bans and quarantining of international arrivals [1]. There was a resurgence of COVID-19 cases in the Australian state of Victoria starting in June, with almost all COVID-19 cases (95%) occurring in people living in the Victoria’s capital Melbourne and surrounding area, a population of 4.93 million out of the total Victorian population of 6.36 million. The resurgence led to the Victorian Government progressively introducing a series of control measures. These measures led to an initial slowing of the growth of the epidemic, then a rapid decline in cases and eventual local epidemic control (i.e., no observed cases for ≥28 days). New diagnoses of COVID-19 in Melbourne peaked at 652 cases on the 4 August 2020. The last diagnosed case associated with this resurgence was reported on the 28 October. A description of this “second wave”, and associated control measure and outcomes has been published [1]. As determined from the sequences of 12,320 SARS-CoV-2 isolates (collected from 1 May 2020 to 30 September 2020) submitted to the GIUSAID database, 97.7% of circulating virus was the D.2 lineage of the GR clade.

The restrictions were primarily applied to metropolitan Melbourne and the adjoining shire of Mitchell (hereafter “Melbourne”), although some were applied state-wide. In Melbourne, from June to October there were five main stages of the restrictions: (1) “pre-Stage 3” (2) “Stage 3”; (3) “Stage 3 + masks”; (4) “Stage 4”; and (5) a gradual easing of restrictions (Table 1). These major directives were accompanied by a vigorous testing of symptomatic people, extensive contact tracing, isolation of infected people and quarantining of close contacts.

**Table 1:** Summary of COVID-19 Restrictions in Melbourne

| **Date:** | **Directions** |
| --- | --- |
| June 21 at 11:59 pm | Relaxation of earlier restrictions enforced during first COVID-19 wave, retaining some restrictions including restrictions on visits to age care and health facilities and on the number of people in private gatherings (5), public gatherings (10) except for weddings (20) and funerals (50). It imposed a one person per 4 sq meter rule for pubs, bars, restaurants etc. [10] |
| July 2 at 11:59 pm | “Stage 3” restrictions to the 10 postcodes in Melbourne with the highest case burden (Stage 3 included: work or study from home where possible: closure of pubs, bars, entertainment venues, churches/places of worship; restricting restaurants and cafes to take-away only, and limiting public gatherings to two people) [11] |
| July 8 at 11:59pm | Extension of Stage 3 restrictions introduced to the rest of Melbourne including the adjoining Mitchell Shire [12, 13] |
| July 20 | Government announces that the mandatory use of masks or face coverings will be introduced in public settings for Melbourne on the July 22 at 11.59 pm [14] |
| July 22 at 11:59pm | Addition of Mandatory use of masks or face coverings in public settings to Stage 3 restrictions for Melbourne [15] |
| August 2 at 6:00 pm | “Stage 4” restrictions for Melbourne (excluding Mitchell Shire). In addition to Stage 3 restrictions, they include: Curfew 8:00 pm to 5:00 am, restriction on movement to <5 Km, substantially increased restrictions on businesses and closure of non-essential stores, no visitor to homes [16, 17] |
| September 13th | Step 1 of eased restrictions. Very minor easing: Curfew changed from 8:00 to 9:00 pm. People living alone can have single visitor allowed in homes, playgrounds re-opened [18, 19] |
| October 18 | Step 2 of eased restrictions. Minor easing: Curfew scrapped, limited opening of schools, some restrictions on non-essential businesses relaxed. Outdoor meetings with 10 people permitted<sup>13</sup> |

A previous regression analysis showed that the introduction of Stage 3 restrictions was associated with a statistically significant reduction in the epidemic growth rate, but it was insufficient to stop the epidemic expansion [2]. Similarly the introduction of mandatory masks, in addition to Stage 3 restrictions, was associated with an exponential but slow decline in cases[3]. Detailed information on the impact of the introduction of masks, including the level of uptake of mask usage and the impact of potentially confounding factors e.g. infections in health care workers, changed movement patterns have been previously reported and therefore are not further considered in this study [3]. The incremental impact of Stage 4 restrictions have not been previously assessed. These two prior analyses considered restricted time-periods over the epidemic wave (from 14 June to 7 July, and from 10 July to 10 August, respectively), with a common theme being that simple exponential models provided excellent fits to the time series of daily detected cases in the first three stages of restrictions (i.e., linear models fitted the logarithm of daily detected cases).

In this analysis, we fit a statistical model to the combined time-series of detected cases across the four restriction phases (pre-Stage 3; Stage 3; Stage 3 + masks and Stage 4) to estimate and compare:

1. the incremental impact of each set of restrictions on the epidemic growth/decline rate. This expands previous analyses by estimation of the potential impact of Stage 4 restrictions and provides better estimates of the epidemic growth/decline in all stages due to the use of a larger data set;
2. the utility of the exponential regression model to describe the daily ranges in observed cases; and
3. the validity of using simple exponential model to predict future trends in daily cases as a tool for planning interventions.

The findings of this work are of considerable interest, as to date Melbourne is one of the few cities globally that experienced a significant outbreak of COVID-19 and then regained control of the epidemic following the introduction of increasing government restrictions and mask use.

## Methods

### Data

Daily diagnosed cases for each Local Government Area (LGA) from 25 Jan 2020 to 20 Nov 2020 were downloaded on the 21 Nov 2020 as the NCOV_COVID_Cases_by_LGA_Source_20201120.cvs file from the Victorian Department of Health and Human Services (DHHS) website [4]. Infections in people living outside of metropolitan Melbourne or acquired overseas were excluded.

### Time-period of interest

The predictive model was based on reported cases a from 14 June to 14 September 2020 inclusive, and hence covers epidemic growth/decline in four distinct phases of restrictions: “pre-Stage 3”, “Stage 3”, “Stage 3 + masks”, and “Stage 4”.

The 14 June starting day was selected to match the previous study on the introduction of “Stage 3” restrictions in Melbourne [2] and because the daily case numbers were inadequate to prevent significant heteroskedasticity at earlier times. On the 14 September, a decision was made to stop the periodic updates of the model. By this date, there was a clear downward trend in daily cases and stopping the analysis on the 14 September allowed testing the ability of the regression model to predict future cases. The primary analysis reported in this paper (Fig. 1A) was conducted on the 14 September 2020.

**Fig. 1.**
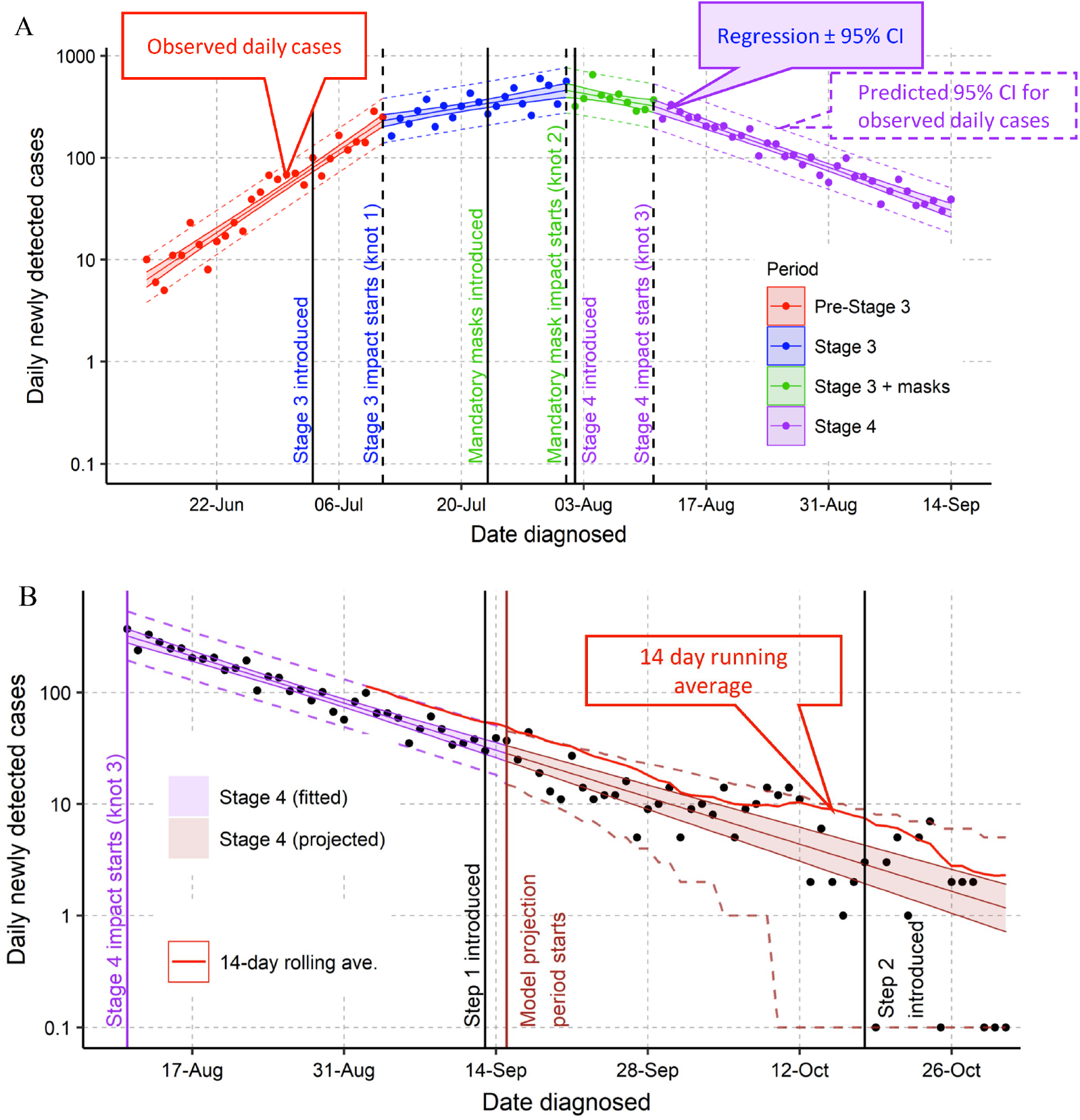
Regression analysis of ln(daily cases) as a function of date of diagnosis for A: the period analysed (14 June to 14 September 2020) and B: extrapolated from the 15 September to 31 October 2020

### Statistical model

The growth or decay rates were estimated using a non-weighted linear regression to fit the natural logarithm of the daily cases against time, using a linear spline model with three knots with the lspline R routine [5]. The three knots divide the model into 4 linked linear segments with the knot days (i.e., the day on which the linear segments are “tied” together) corresponding to the times at which impact of successively introduced control procedures are predicted to start. The knots allow deflections in the line of best fit due to the impact of the introduction of different restrictions. The assumption of linearity of the four segments, and hence the presumption of an exponential growth/decay was tested by assessing whether studentized residuals were normally distributed and independent of the time (i.e., no significant heteroskedasticity).

Due to the generation interval for SARS-CoV-2 infection (a mean of 4-6 days [6, 7]) and delays in testing and reporting, there is an expected delay between the introduction of restrictions and observing them in diagnosis data. The knot days were estimated to be 8 days following the introduction of new control measures; for example, the introduction of Stage 3 restrictions took place on 2 July, so the knot day was 3 July + 8 days = 11 July. This delay was based on a previous study that estimated an 8-day delay between the introduction of mandatory face masks in Melbourne and the change in the epidemic growth rate [3].

This analysis estimated the instantaneous growth constant (*k*) (i.e., the slope of the ln(daily cases) vs time regression) for each segment and the difference between these growth constants was used as the primary measure to test for a significant association between successive public health measures being introduced and subsequent changes in growth/decay rates.

The doubling/halving time of the epidemic was calculated as *t*_2_ = ln(2)/*k*

### Effective Reproduction ratio (R_eff_)

We calculated *R*_*eff*_ as the ratio of new cases separated by one serial interval for SARS-CoV-2 infections using the Lotka–Euler equation [8]. Due to uncertainties in the serial interval, we estimate a range for *R*_*eff*_ based on two measurements of the serial interval: a normal distribution with mean 3.96, standard deviation 4.75 days based on 468 pairs of infections in China [6], and a gamma distribution with mean 6.49, standard deviation 4.90 days based on 1015 pairs of infections in China, Japan and Singapore [7]. We include changes in estimated *R*_*eff*_ as a secondary estimate to allow comparisons with other studies but emphasize that the estimates of *R*_*eff*_ are subject to uncertainty in estimates of the generation interval (approximated as the serial interval) and its distribution.

We also use the change in *R*_*eff*_ as a measure of the relative impact of successively increasing control measures for both the 3.96 and the 6.49-day serial interval estimates.

### Extension to 31 October

To test the utility of the regression analysis to predict future trends, we extrapolated the regression line from the last day of data fitted (14 September) to the end of October. We calculated the predicted 95% range of daily cases in this period assuming an inverse Poisson distribution using two methods: the estimated daily cases calculated as the antilog of the mean ln(cases) or the mean ln(cases) ±the 95% confidence intervals from the regression line, to provide minimum and maximum estimates of the range, respectively.

## Results

### Regression analysis 14 July to 14 September2020

The regression analysis of the ln(daily cases) vs time is shown in Fig. 1A for the model with four log-linear segments joined by three knots with each knot 8 days after the introduction of the increased control measures (Fig. 1). For each log-linear segment, the corresponding instantaneous growth rate, doubling/having time and *R*_*eff*_ are shown are shown in Table 2, alongside the percentage reduction in *R*_*eff*_ between segments. These growth constants translate into from a 14% daily increase in pre-stage 3 to a 3.3 % increase in daily cases during of Stage 3; then to a 3.5% decrease in daily cases after introduction of mandatory masks and a further a 6.7% decrease in Stage 4.

**Table 2.** Growth Constants, doubling times and Reff for each stage.

| Control Level | Growth constant per day | $p > 0$ | $p k_n - k_{n-1} > 0$ | Doubling/halving time (95% CI) days | Serial Interval 3.96 | Successive Reduction | Serial Interval 6.49 | Successive Reduction |
| --- | --- | --- | --- | --- | --- | --- | --- | --- |
| | | | | | $R_{eff}$ (95% C.I.) | | $R_{eff}$ (95% C.I.) | |
| Pre-Stage 3 | 0.133 (s.e. 0.005) | <0.001 | - | 5.22 (4.87 to 5.61) | 1.39 (1.37 to 1.40) | - | 2.02 (1.93 to 2.1) | - |
| Stage 3 | 0.033 (s.e. 0.006) | <0.001 | <0.001 | 21.21 (15.75 to 32.49) | 1.12 (1.08 to 1.16) | 18.5% | 1.22 (1.14 to 1.3) | 35.4% |
| Stage3 + masks | -0.036 (s.e. 0.012) | 0.003 | <0.001 | -19.47 (-54.63 to -11.84) | 0.86 (0.76 to 0.95) | 25.9% | 0.78 (0.65 to 0.92) | 39.0% |
| Stage 4 | -0.069 (s.e. 0.004) | <0.001 | 0.019 | -10 (-11.19 to -9.03) | 0.72 (0.69 to 0.75) | 14.7% | 0.59 (0.56 to 0.63) | 22.6% |

Intercept: 1.86 s.e. 0.086; Residual standard error: 0.24 on 88 degrees of freedom; Multiple R-squared: 0.959 Adjusted R-squared: 0.957; F-statistic: 509 on 4 and 88 DF, p-value: < 0.001. Doubling/halving time calculated as $\ln(2)/k$ (see text)

Over the whole of the regression, the observed daily cases were only outside the 95% predicted confidence limits for three of the 93 days analysed. I.e., the distribution fitted on 96% of the days as expected for a 95% confidence interval.

The 4-segment linear exponential model gave a good fit of the observed data to the model as judged by the goodness-of fit tests detailed in the Supplementary data, but to summarize: the distribution of studentized residual values of the log transformed cases was indistinguishable from a normal distribution (QQ plot and Shapiro-Wilk Test, p= 0.73). There was no significant autocorrelation (Durbin-Watson Test) and no outlier data that significantly impacted on the regression (maximum Cook’s D value was 0.085). However, one test that showed a deviation from an ideal regression: there was evidence of heteroskedasticity (Breusch-Pagan test p= 0.0013, White test p=0.0014). Inspection of the residuals indicted that the initial data points had a higher scatter consistent with the lower daily cases in June. A regression using three segments with two knots for the period 11 July to 14 September (i.e., starting at the day of the first expected impact of Stage 3) gave a fit with no significant heteroskedasticity and the growth constants for the stage 3, stage 3+masks and stage 4 were essentially unchanged from those obtained with the full model (Table 2). Thus, the observed heteroskedasticity at in the start of the first segment had no impact on the estimates of growth constants for stage 3, stage 3+masks or Stage 4.

### Extrapolation to the 31 October 2020

The observed distribution of cases over the period 15 September to 31 October closely matched the projected distribution, with most cases lying within the 95% confidence intervals. Importantly, this projection showed a high probability of zero cases at the end of October (31% probability of zero cases on the 31 October). Since the confidence interval for the regression line was small compared to the Poisson confidence intervals, there was little difference in the confidence interval calculated with the two methods based on the mean or the mean ± 95% confidence interval.

## Discussion

From 14 June to 14 September, the four-segment linear model gave a good fit to the observed daily cases in Melbourne. There was a highly significant change in the growth constants (and associated *R*_*eff*_) following the three major changes in control measures. The biggest successive impact as measured by the successive percent change in *R*_*eff*_, was the addition of masks to the existing Stage 3 restrictions, followed by increased restrictions at the start Stage 3, regardless of the underlying assumptions about serial interval (Table 2).

Although introduction of masks had the largest effect, the addition of further restrictions at the start of Stage 4 still was associated with a statistically significant change in the growth constant. Without Stage 4, the extrapolated trend predicted 18 new cases per day on the 31 October compared to the 1.2 cases predicted with the additional Stage 4 restrictions. Alternatively, it would have taken until the 15 January to reach an average of 1.2 cases per day. Also, the addition of Stage 4 is estimated to have prevented approximately 3900 cases compared to continued use of just Stage 3 + masks until local epidemic control.

Growth constants and associated *R*_*eff*_ had previously been reported using regression analyses for the initial growth period and Stage 3 for all cases in Victoria [2, 3] and for Stage 3 with and without masks for Melbourne cases [3]. The analyses in this paper used a wider range of data and importantly, included the additional Stage 4 restrictions: The first study fitted the data to two independent linear regression lines of the ln(daily cases) vs time and the second to a model with two linear segments and a single knot. As a result of the larger database and by constraining the end of each segment through the use of the knots, the precision of the estimates are improved as judged by the smaller standard errors of the estimated growth constants in the current study compared to the earlier studies (Table 3).

**Table 3.** Comparison of coefficients from previous studies with the current study

|  | Pre-Stage 3 |  |  | Stage 3 |  |  | Stage 3+ Masks |  |  |
| --- | --- | --- | --- | --- | --- | --- | --- | --- | --- |
| | Growth Constant | Std. Error | $R_{eff}$ | Growth Constant | Std. Error | $R_{eff}$ | Growth Constant | Std. Error | $R_{eff}$ |
| Victoria<sup>2</sup> | 0.14 | 0.008 | 1.74 | 0.038 | 0.009 | 1.16 |  |  |  |
| Melbourne<sup>3</sup> |  |  |  | 0.042 | 0.007 | 1.18 | -0.023 | 0.017 | 0.91 |
| Melbourne, this study | 0.133 | 0.005 | 1.39 | 0.033 | 0.006 | 1.12 | -0.036 | 0.012 | 0.86 |

\* $R_{eff}$ Calculated assuming a 3.96 day serial interval

As previously noted, earlier work considered a range of possible confounding factors that might change the estimated growth rates in the Stage 3 to Stage 3 + Masks transition [3], including the impact of COVID-19 cases on testing patterns, infections in health care workers and the impact of changed (reduced) movement patterns. None of the potential confounding factors had a significant impact on the estimated changes in the growth constant going from Stage 3 to Stage 3 + Masks and were not considered in this broader study. However, as concluded in the previous work, although no statistically significant impact was observed, these confounding factors were likely to result in an under-estimate of the impact of each of the increased restrictions. For example, health care workers were already using masks and other PPE over the whole period.

Our results have several implications for real time control decisions to guide the future imposition or relaxation of control measures. These include:

1. Managing community expectation in the fluctuations of observed daily cases. The power of the statistics associated with a regression analysis provides an estimate of the expected daily range. As shown in Fig 1, this analysis accurately predicted the range of daily cases expected (i.e., for the main analysis period, for 90 out of 93 days (96%), the observed distribution lay within the 95% estimated range). It will give a short-term estimate of expected range. If applied in future outbreaks may give either reassurance or a rapid warning that the epidemic situation is changing.
2. Longer term prediction of future trends. Extrapolation of the regression analysis from the 14 September to the 31 October correctly predicted the trend of the epidemic. This approach differs from the 14-day rolling average approach used in Victoria to provide Go/No Go decision on relaxing controls [1]. Because the regression analysis is based on all available data and built around a statistical model, it can be projected into the future with realistic confidence ranges. The rolling average is based on much less data (14 days) and is subject to greater variation (Fig. 1B) and gives an historical view on where the epidemic was on average 7 days previously. It cannot provide estimates of the expected daily fluctuations in case number or a statistical basis including confidence ranges on predicting future cases.
3. Quantitative data on the impact of progressively imposed control measures. As judged by *R*_*eff*_, this study highlights that the single most important sequential control measure was the addition of mandatory masks. The much more stringent control measures subsequently imposed with Stage 4 had a significant but smaller impact on the time to epidemic control but at a much greater cost to the community. If future, with a much smaller outbreak, Stage 3 + masks alone may be sufficient. Alternatively, if faced with a more highly transmissible strain [9] the data from this study may be useful in calibrating future models of what will be required for local epidemic control.

## Conclusions

A simple and easily calculated linear regression model with knots at each significant change in control measures, gave an adequate fit to the observed daily cases in Melbourne’s second COVID-19 wave, providing quantitative data on the relatively impact of the different controls and provides a framework for simple models to guide future outbreaks responses should this be required.

## Supporting information

Goodness of fit for regression

## Data Availability

All data are available publicly from sources cited in the MS. Specifically, from the Victorian Government site

https://www.dhhs.vic.gov.au/ncov-covid-cases-by-lga-source-csv

## Abbreviations

DHHS: Victorian Department of Health and Human Services
*k*: instantaneous growth constant
*R*_*eff*_: Effective Reproduction Ratio
s.e.: standard error
C.I.: confidence interval

## Acknowledgements

We acknowledge access to the sequence data of SARS-CoV-2 isolates in the GISAID database (https://www.gisaid.org/) submitted by the Microbiological Diagnostic Unit - Public Health Laboratory of The Peter Doherty Institute for Infection and Immunity, 792 Elizabeth Street, Melbourne, Victoria, Australia, 3000.

## Supplementary Data

Supplement.pdf: goodness of fit tests for the regression analysis

