## Supplementary material for "The impact of three progressively introduced interventions on second wave daily COVID-19 case numbers in Melbourne, Australia": Goodness of fit for regression

### A. Goodness of fit tests for regression analysis

This supplement describes the tests undertaken on the regression analysis shown in Figure 1A of the main paper.

In a valid multiple linear regression, it is expected that the residuals meet the following criteria:

- **Linearity**: The $\varepsilon_i$ have mean of 0
- **Independence**: The $\varepsilon_i$ are independent
- **Normality**: The $\varepsilon_i$ are normally distributed
- **Homogeneity of variances**: The $\varepsilon_i$ have the same variance $\sigma^2$

We primarily test using the Studentized residuals $\mathcal{E}_i$ since raw residuals are not expected to be completely independent but provide some test results with the raw residuals

**Linearity**

| | Mean |
|---|---|
| Raw Residuals | $4.0 \times 10^{-16}$ |

Expect zero. Pass

**Independence**

*Durban-Watson Test*

| | |
|---|---|
| Test value negative autocorrelation | 2.040 |
| Test value postitive autocorrelation | 1.574 |
| Critical upper limit for α = 0.05 | 1.753 |

Test values are both greater than the critical upper limit so no positive or negative autocorrelation is detected.  Therefore there is no evidence for lack of independence.

*Graphical display*

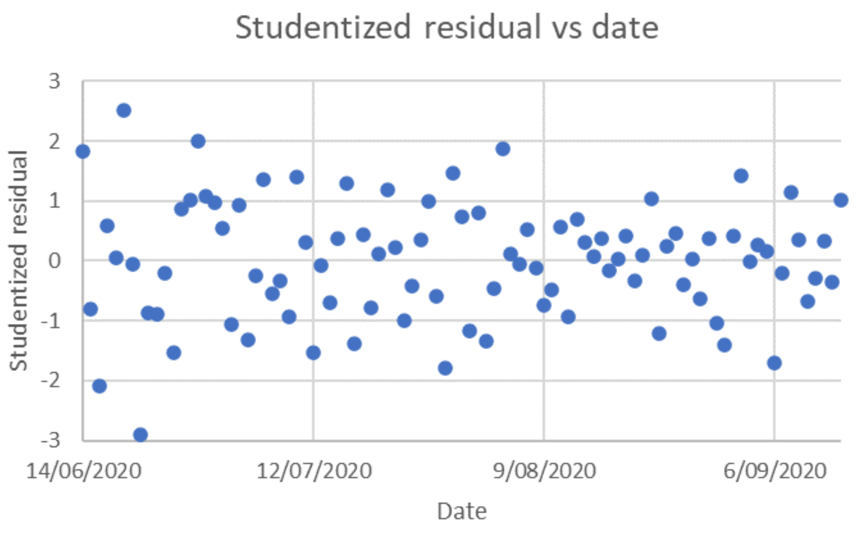

Fig. S1. Plot of Studentized residuals *versus* day of the study. The knot days were 11 July, 1 August and 12 August

**Normality**

*Shapiro-Wilk test*

| | residual |
|---|---|
| W-stat | 0.997 |
| p-value | 0.8321 |

Test passes for residuals ($p>0.05$)

*Graphical QQ Plot*

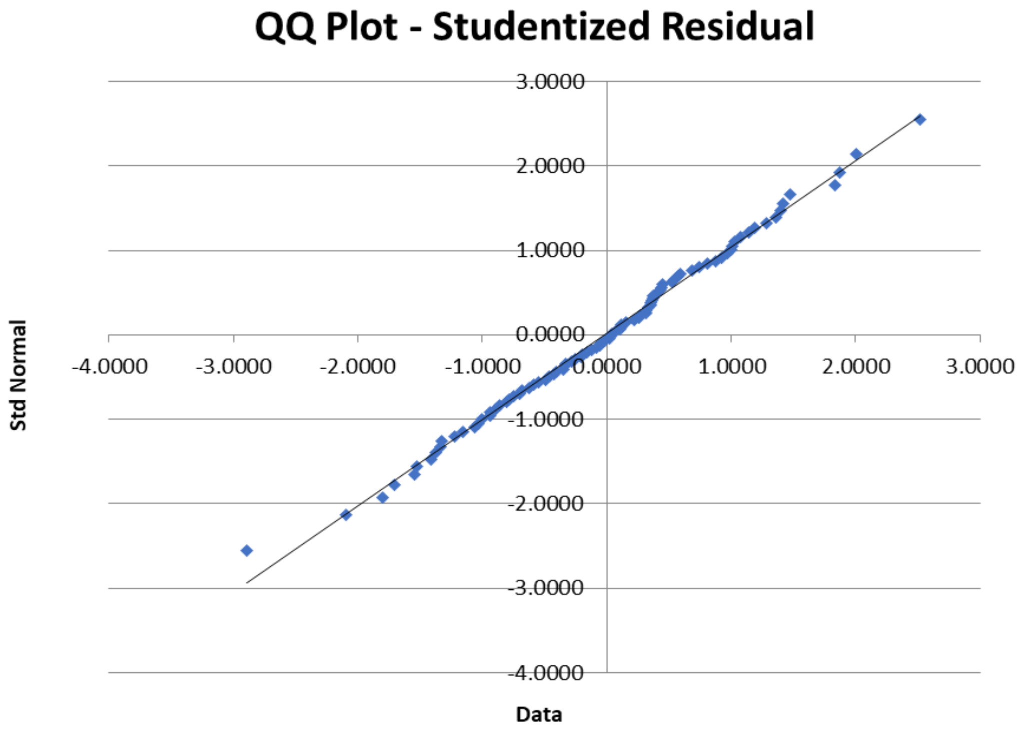


Fig. S2. QQ (Quantile-Quantile) plot of the ordered Studentized residuals vs a the corresponding quantiles of a normal distribution. Black line indicates the position of a perfectly normally distributed Studentized residuals.

**Homogeneity of Variances**

*Test for Heteroskedasticity with Breusch-Pagan test over the range 14 June to 14 September (i.e pre-Stage 3 to the end of the analysis in Stage 4)*

| | |
|---|---|
| Number of Days | 93 |
| No. of Independent variables | 4 |
| LM statistic | 17.81 |
| Degrees of Freedom | 2 |

| | |
|---|---|
| p-value | 0.0013 |

Pass criteria: p value >0.05.  Fail

*Test for Heteroskedasticity with Breusch-Pagan test over the range 11 July to 14 September (i.e just Stage 3 to the end of the analysis in Stage 4)*

| | |
|---|---|
| Number of Days | 66 |
| No. of Independent variables | 4 |
| LM statistic | 7.57 |
| Degrees of Freedom | 2 |
| p-value | 0.11 |

Pass criteria: p value >0.05.  pass

Also see Fig S1

**Other test**

Regression was tested for the infludence of outliers using Cook's D test.  As shown below, the maximum Cook's D value was 0.109 on 16 June 2020  As this is much less than 1, there are no outliers with a significant impact on the regression

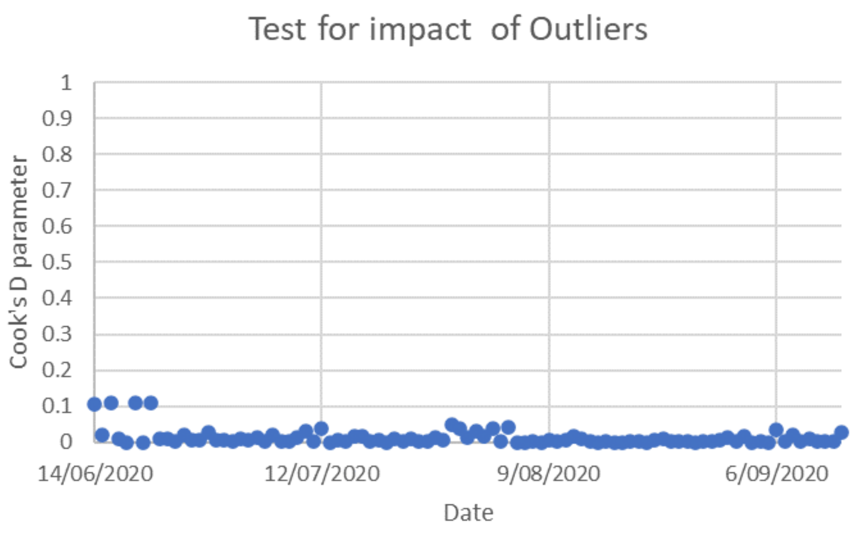
